# Perspectives of Cancer Patients and Their Health during the COVID-19 Pandemic

**DOI:** 10.1101/2020.04.30.20086652

**Authors:** Emil Lou, Deanna Teoh, Katherine Brown, Anne Blaes, Shernan G. Holtan, Patricia Jewett, Helen Parsons, E. Waruiru Mburu, Lauren Thomaier, Jane Yuet Ching Hui, Heather H. Nelson, Rachel I. Vogel

## Abstract

**Introduction:** The immunosuppressive nature of some cancers and many cancer-directed treatments may increase the risk of infection with and severe sequelae from Coronavirus Disease 2019 (COVID-19). The objective of this study was to compare concerns about COVID-19 among individuals undergoing cancer treatment to those with a history of cancer not currently receiving therapy and to those without a cancer history.

**Methods:** We conducted a cross-sectional anonymous online survey study of adults currently residing in the United States. Participants were recruited over a one-week period (April 3-11, 2020) using promoted advertisements on Facebook and Twitter. Groups were compared using chi-squared tests, Fisher’s exact tests, and t-tests.

**Results:** 543 respondents from 47 states provided information on their cancer history and were included in analyses. Participants receiving active treatment reported greater concern about coronavirus infection (p<0.0001), higher levels of family distress caused by the COVID-19 pandemic (p=0.004), and greater concern that the general public does not adequately understand the seriousness of COVID-19 (p=0.04). Those with metastatic disease were more likely to indicate that COVID-19 had negatively affected their cancer care compared to patients with non-metastatic cancer (50.8% vs. 31.0%; p=0.02). The most commonly reported treatment modifications included chemotherapy delays.

**Conclusions:** Patients undergoing active treatment for cancer were most concerned about the short-term effects of the COVID-19 pandemic on the logistics as well as potential efficacy of ongoing cancer treatment, longer term effects, and overarching societal concerns that the population at large is not as concerned about the public health implications of the coronavirus.

## INTRODUCTION

The coronavirus disease 2019 (COVID-19) pandemic has led to sudden shifts in healthcare, including re-categorization of “essential” care^1^. The population of patients with current and previous history of cancer represents a unique subset that as a whole may be more susceptible to coronavirus infection and sequelae of COVID-19^2^. This is thought to be due to the immunosuppressive nature of some cancers and many cancer-directed treatments, and the relatively higher frequency clinical visits for patients undergoing active treatment and for those in active surveillance following treatment compared to patients without cancer^3,4^.

Under usual circumstances, a cancer diagnosis elicits anxiety regarding scheduling logistics, outcomes, and side-effects of cancer-directed treatments^5^. The recent changes in cancer care due to the COVID-19 pandemic^6^, including treatment delays, cancellation of procedures deemed not essential, and a transition into virtual rather than in-person clinic visits, have likely added to these usual uncertainties and fears associated with having cancer.

We conducted a cross-sectional survey study to examine emotional well-being and health care decision-making in individuals with and without a history of cancer. We hypothesized that while distress and anxiety is increased in the general population related to the pandemic, these emotions, would be higher in patients with a diagnosis of cancer. The primary objective of this survey study was to compare emotional well-being and decision-making among cancer patients undergoing therapy during the COVID-19 health crisis to two control groups: 1) cancer survivors who are not currently undergoing treatment and 2) those without a history of cancer.

## METHODS

This anonymous cross-sectional online survey study was approved by the University of Minnesota Institutional Review Board. To be eligible, participants had to be 18 years of age or older, currently reside in the United States, and able to read and write in English. Individuals were recruited over a one-week period (April 3-11, 2020) using promoted ads on the social media apps Facebook and Twitter, with separate ads specifically targeting those with a history of cancer. Invitations to participate in the survey were also posted by the American Cancer Society. Survey data were collected and stored using REDCap, a web-based data collection tool^7^.

Survey items included demographics, personal concerns about COVID-19, and among those currently receiving cancer therapy, perceived effects of the COVID-19 pandemic on cancer treatment. Validated measures were used or modified as appropriate when possible. Symptoms of generalized anxiety and depression were measured using the General Anxiety Disorder-7 (GAD-7)^8^ and Patient Health Questionnaire (PHQ-8)^9^, respectively; potentially clinically relevant cutoffs of 10 or greater were used for each. The number of COVID-19 cases in each state was determined using data from the Centers of Disease Control and Prevention (CDC) as of April 3, 2020^10^.

We used chi-squared tests and t-tests to compare demographic characteristics between the three groups of interest: 1) cancer patients currently undergoing therapy, 2) individuals with a current or past diagnosis of cancer not currently undergoing therapy, 3) individuals with no history of cancer. Descriptive statistics summarized rates of treatment plan changes among cancer patients currently receiving therapy and compared those with and without metastatic disease using chi-squared and Fisher’s exact tests.

## RESULTS

Among the 839 individuals who opened the survey, 815 (97%) were eligible for the study. A total of 543 participants provided information on their cancer history and were included in this analysis. Among eligible participants, 25.6% were undergoing active cancer treatment, 29.8% reported a history of cancer but not on active treatment, and 44.6% reported no history of cancer. Forty-seven states were represented; there were no respondents from Alaska, Vermont or Wyoming. Minnesota had the highest percentage of respondents (24.0%). Only two respondents reported being diagnosed with coronavirus. The average age of participants was 55.5 years (SD=14.4) and the majority were female (82.8%) and non-Hispanic White (95.0%), with 23.9% being responsible for children under 18 years old and 15.2% caring for other adult(s). Those with a history of cancer were older (p=0.001), more likely to be retired (p=0.003) and less likely to have a child under 18 years old living with them (p=0.006; **Table 1**). Participants with cancer currently undergoing treatment were more likely to report their general health as fair or poor (p<0.0001). All other demographic characteristics were generally balanced between the groups, including the number of COVID-19 cases in residing state. Among those ever diagnosed with cancer, the three most common cancers were breast (40.5%), colon (16.6%), and non-melanoma skin cancer (10%). Approximately one-quarter (25.3%) of those with a cancer diagnosis reported stage IV cancer.

**Table 1.**
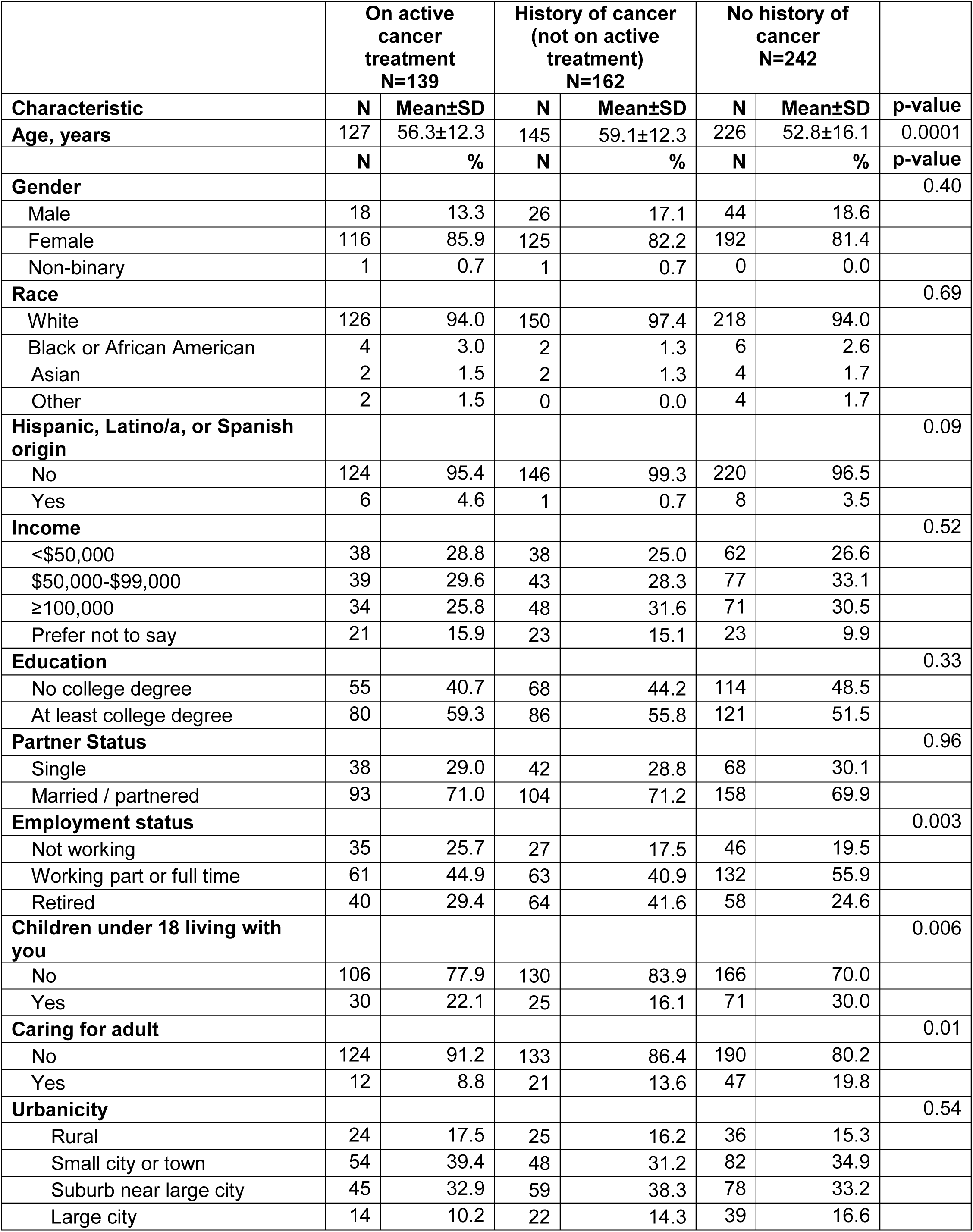
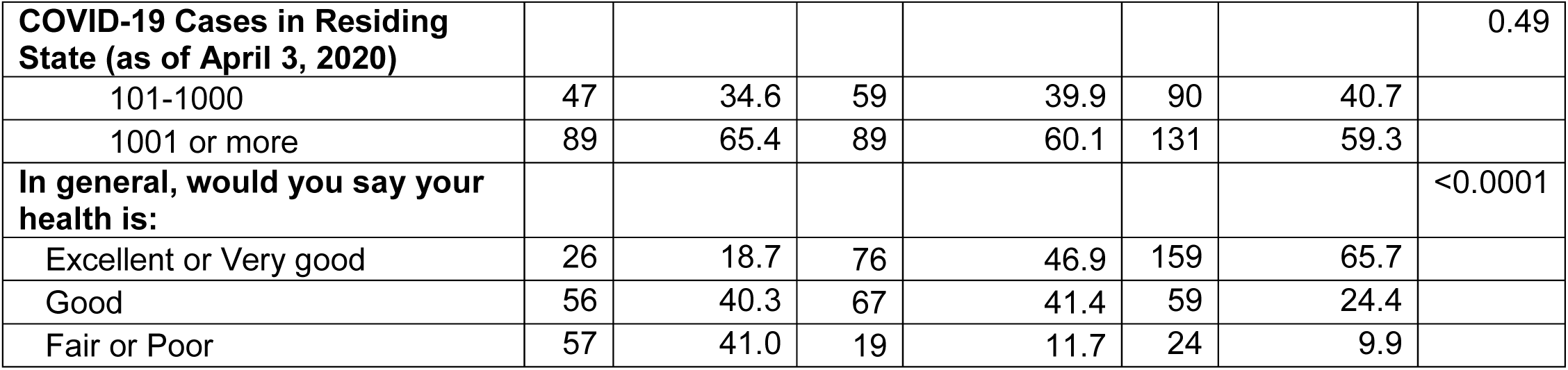
Demographic characteristics.

|  | On active cancer treatment<br>N=139 |  | History of cancer<br>(not on active treatment)<br>N=162 |  | No history of cancer<br>N=242 |  |  |
| --- | --- | --- | --- | --- | --- | --- | --- |
| Characteristic | N | Mean±SD | N | Mean±SD | N | Mean±SD | p-value |
| Age, years | 127 | 56.3±12.3 | 145 | 59.1±12.3 | 226 | 52.8±16.1 | 0.0001 |
|  | N | % | N | % | N | % | p-value |
| Gender |  |  |  |  |  |  | 0.40 |
| Male | 18 | 13.3 | 26 | 17.1 | 44 | 18.6 |  |
| Female | 116 | 85.9 | 125 | 82.2 | 192 | 81.4 |  |
| Non-binary | 1 | 0.7 | 1 | 0.7 | 0 | 0.0 |  |
| Race |  |  |  |  |  |  | 0.69 |
| White | 126 | 94.0 | 150 | 97.4 | 218 | 94.0 |  |
| Black or African American | 4 | 3.0 | 2 | 1.3 | 6 | 2.6 |  |
| Asian | 2 | 1.5 | 2 | 1.3 | 4 | 1.7 |  |
| Other | 2 | 1.5 | 0 | 0.0 | 4 | 1.7 |  |
| Hispanic, Latino/a, or Spanish origin |  |  |  |  |  |  | 0.09 |
| No | 124 | 95.4 | 146 | 99.3 | 220 | 96.5 |  |
| Yes | 6 | 4.6 | 1 | 0.7 | 8 | 3.5 |  |
| Income |  |  |  |  |  |  | 0.52 |
| <\$50,000 | 38 | 28.8 | 38 | 25.0 | 62 | 26.6 | |
| \$50,000-\$99,000 | 39 | 29.6 | 43 | 28.3 | 77 | 33.1 | |
| ≥100,000 | 34 | 25.8 | 48 | 31.6 | 71 | 30.5 |  |
| Prefer not to say | 21 | 15.9 | 23 | 15.1 | 23 | 9.9 |  |
| Education |  |  |  |  |  |  | 0.33 |
| No college degree | 55 | 40.7 | 68 | 44.2 | 114 | 48.5 |  |
| At least college degree | 80 | 59.3 | 86 | 55.8 | 121 | 51.5 |  |
| Partner Status |  |  |  |  |  |  | 0.96 |
| Single | 38 | 29.0 | 42 | 28.8 | 68 | 30.1 |  |
| Married / partnered | 93 | 71.0 | 104 | 71.2 | 158 | 69.9 |  |
| Employment status |  |  |  |  |  |  | 0.003 |
| Not working | 35 | 25.7 | 27 | 17.5 | 46 | 19.5 |  |
| Working part or full time | 61 | 44.9 | 63 | 40.9 | 132 | 55.9 |  |
| Retired | 40 | 29.4 | 64 | 41.6 | 58 | 24.6 |  |
| Children under 18 living with you |  |  |  |  |  |  | 0.006 |
| No | 106 | 77.9 | 130 | 83.9 | 166 | 70.0 |  |
| Yes | 30 | 22.1 | 25 | 16.1 | 71 | 30.0 |  |
| Caring for adult |  |  |  |  |  |  | 0.01 |
| No | 124 | 91.2 | 133 | 86.4 | 190 | 80.2 |  |
| Yes | 12 | 8.8 | 21 | 13.6 | 47 | 19.8 |  |
| Urbanicity |  |  |  |  |  |  | 0.54 |
| Rural | 24 | 17.5 | 25 | 16.2 | 36 | 15.3 |  |
| Small city or town | 54 | 39.4 | 48 | 31.2 | 82 | 34.9 |  |
| Suburb near large city | 45 | 32.9 | 59 | 38.3 | 78 | 33.2 |  |
| Large city | 14 | 10.2 | 22 | 14.3 | 39 | 16.6 |  |
| **COVID-19 Cases in Residing State (as of April 3, 2020)** |  |  |  |  |  |  | 0.49 |
| 101-1000 | 47 | 34.6 | 59 | 39.9 | 90 | 40.7 |  |
| 1001 or more | 89 | 65.4 | 89 | 60.1 | 131 | 59.3 |  |
| **In general, would you say your health is:** |  |  |  |  |  |  | <0.0001 |
| Excellent or Very good | 26 | 18.7 | 76 | 46.9 | 159 | 65.7 |  |
| Good | 56 | 40.3 | 67 | 41.4 | 59 | 24.4 |  |
| Fair or Poor | 57 | 41.0 | 19 | 11.7 | 24 | 9.9 |  |

Rates of generalized anxiety and depression were similar across groups (**Table 2**). However, those in the active treatment group were more likely to report high levels of concern about getting infected by SARS-CoV-2 (71.9%), which was significantly higher than those who had completed cancer treatment (47.9%) or had no history of cancer (51.9%; p<0.0001). In contrast, there was a gradient across groups when asked about their risk for a severe manifestation of the infection (p<0.0001); only 33.8% of those with no cancer history felt they were at high risk for severe disease compared with 58.0% of those who had completed cancer treatment, and 90.7% of those who were in active treatment. Most respondents in the three groups reported that they regarded COVID-19 as a “moderately” or “very” serious threat, but patients with a history of previously treated cancer or cancer undergoing active treatment were more likely to report practicing complete social/physical distancing in the previous week as compared to individuals without cancer (p=0.0009). Despite this difference, a majority of all three groups (>80%) reported high concern about close family members or friends becoming infected. Patients undergoing active treatment reported the highest level of family distress caused by the COVID-19 pandemic (p=0.004), as well as concern that some others did not adequately understand the seriousness of COVID-19 and its implications (p=0.04).

**Table 2.** Emotional health and COVID-19 experience and concerns.

| Variable | On active cancer treatment<br>N=139 |  | History of cancer<br>(not on active treatment)<br>N=162 |  | No history of cancer<br>N=242 |  | p-value |
| --- | --- | --- | --- | --- | --- | --- | --- |
|  | N | % | N | % | N | % |  |
| **Generalized Anxiety** |  |  |  |  |  |  | 0.49 |
| No | 99 | 73.9 | 122 | 78.7 | 174 | 73.7 |  |
| Yes | 35 | 26.1 | 33 | 21.3 | 62 | 26.3 |  |
| **Depression** |  |  |  |  |  |  | 0.07 |
| No | 96 | 73.3 | 125 | 82.2 | 171 | 72.5 |  |
| Yes | 35 | 26.7 | 27 | 17.8 | 65 | 27.5 |  |
| **How concerned are about getting COVID-19?** |  |  |  |  |  |  | <0.0001 |
| Not at all or slightly concerned | 15 | 11.1 | 31 | 19.6 | 64 | 26.7 |  |
| Somewhat concerned | 23 | 17.0 | 45 | 28.5 | 61 | 25.4 |  |
| Moderately or extremely concerned | 97 | 71.9 | 82 | 51.9 | 115 | 47.9 |  |
| **Consider yourself to be at "high risk" for severe illness from COVID-19?** |  |  |  |  |  |  | <0.0001 |
| No | 7 | 5.0 | 32 | 19.8 | 120 | 50.0 |  |
| Yes | 126 | 90.7 | 94 | 58.0 | 81 | 33.8 |  |
| Unsure | 6 | 4.3 | 36 | 22.2 | 39 | 16.3 |  |
| **How serious do you think COVID-19 is?** |  |  |  |  |  |  | 0.07 |
| Not at all or a little serious | 2 | 1.4 | 1 | 0.6 | 8 | 3.3 |  |
| Somewhat serious | 2 | 1.4 | 5 | 3.1 | 14 | 5.8 |  |
| Moderately or very serious | 135 | 97.1 | 156 | 96.3 | 220 | 90.9 |  |
| **How much have you been social distancing in the past week?** |  |  |  |  |  |  | 0.0007 |
| Not at all or a little | 2 | 1.4 | 0 | 0.0 | 7 | 2.9 |  |
| Some or mostly | 42 | 30.2 | 56 | 34.6 | 111 | 45.9 |  |
| Completely | 95 | 68.4 | 106 | 65.4 | 124 | 51.2 |  |
| **How concerned about one of your close family members or friends getting COVID-19?** |  |  |  |  |  |  | 0.07 |
| Not at all or slightly concerned | 12 | 8.6 | 4 | 2.5 | 22 | 9.1 |  |
| Somewhat concerned | 15 | 10.8 | 15 | 9.3 | 23 | 9.5 |  |
| Moderately or extremely concerned | 112 | 80.6 | 143 | 88.3 | 197 | 81.4 |  |
| **How concerned about getting needed healthcare if you become seriously ill from COVID-19?** |  |  |  |  |  |  | 0.99 |
| Not at all or slightly concerned | 25 | 18.0 | 27 | 16.7 | 45 | 18.6 |  |
| Somewhat concerned | 26 | 18.7 | 29 | 17.9 | 44 | 18.2 |  |
| Moderately or extremely concerned | 88 | 63.3 | 106 | 65.4 | 153 | 63.2 |  |
| **How concerned about getting needed healthcare if you become ill from something else?** |  |  |  |  |  |  | 0.75 |
| Not at all or slightly concerned | 32 | 23.0 | 36 | 22.2 | 65 | 26.9 |  |
| Somewhat concerned | 31 | 22.3 | 31 | 19.1 | 47 | 19.4 |  |
| Moderately or extremely concerned | 76 | 54.7 | 95 | 58.6 | 130 | 53.7 |  |
|  | **N** | **Mean±SD** | **N** | **Mean±SD** | **N** | **Mean±SD** | **p-value** |
| **How distressing has COVID-19 been for your family?** | 136 | 74.5±19.3 | 157 | 69.8±21.5 | 240 | 66.7±23.8 | 0.004 |
| **Interference of COVID-19 with (self-) employment** | 133 | 36.5±37.0 | 151 | 34.1±36.5 | 236 | 40.1±37.1 | 0.27 |
| **Interference of COVID-19 with activities at home** | 137 | 52.5±31.0 | 154 | 52.6±30.1 | 231 | 52.8±30.4 | 0.99 |
| **Isolation related to COVID-19?** | 138 | 74.0±26.5 | 156 | 68.0±28.7 | 236 | 67.7±26.5 | 0.08 |
| **Financial burden from COVID-19 to date** | 135 | 41.4±33.3 | 152 | 38.4±34.2 | 237 | 40.4±33.2 | 0.73 |
| **Financial burden from COVID-19 over next 12 months** | 138 | 53.6±31.4 | 154 | 51.0±33.0 | 237 | 54.2±33.0 | 0.62 |
| **How worried about paying for medical care during the COVID-19 pandemic?** | 137 | 36.6±34.7 | 151 | 31.5±33.9 | 233 | 36.4±33.7 | 0.33 |
| **How concerned about some people not understanding the seriousness of COVID-19?** | 137 | 89.0±16.1 | 158 | 83.9±21.3 | 234 | 83.9±21.7 | 0.04 |

Among those undergoing therapy, 20.7% reported no contact with their oncologists about treatment plans since the pandemic began. Those with metastatic cancer were significantly more likely to have had contact with their oncologist than those with non-metastatic disease (29.2% vs. 13.1%; p=0.03) (**Table 3**). A higher proportion of participants with metastatic cancer reported that they “strongly” or “somewhat strongly” agreed with the statement that COVID-19 had negatively affected their cancer care, compared to participants with non-metastatic cancer (50.8% vs. 31.0%; p=0.02). Some (17.9%) of those currently receiving cancer therapy stated the pandemic changed their ongoing treatment plans. While this did not differ between those with and without metastatic disease, those who did report having treatment plans changed were more likely to report clinically relevant symptoms of anxiety (52.0% vs. 17.0%, p=0.0003) and depression (56.0% vs. 20.0%, p=0.0004) than those who did not report a treatment plan change.

**Table 3.** Comparison of cancer treatment concerns among those currently undergoing therapy or diagnosed with cancer during COVID-19 by cancer stage (metastatic vs. non-metastatic; N=133).

|  | Non-metastatic |  | Metastatic |  |  |
| --- | --- | --- | --- | --- | --- |
| Variable | N | % | N | % | p-value |
| **In contact with oncologist about treatment plan since COVID-19 pandemic began** |  |  |  |  | 0.03 |
| No | 21 | 29.2 | 8 | 13.1 |  |
| Yes | 51 | 70.8 | 53 | 86.9 |  |
| **The COVID-19 pandemic has negatively affected my cancer care.** |  |  |  |  | 0.02 |
| Strongly or somewhat agree | 22 | 31.0 | 31 | 50.8 |  |
| Neutral/Somewhat agree/Strongly disagree | 49 | 69.0 | 30 | 49.2 |  |
| **Were your appointments moved to telephone visits or video encounters (telehealth)?** |  |  |  |  | 0.59 |
| Yes telephone always | 33 | 64.7 | 28 | 52.8 |  |
| Yes telephone sometimes | 2 | 3.9 | 1 | 1.9 |  |
| Yes video visits always | 8 | 15.7 | 10 | 18.9 |  |
| Yes video visits sometimes | 2 | 3.9 | 5 | 9.4 |  |
| Yes combination of telephone and video visits | 1 | 2.0 | 4 | 7.6 |  |
| No | 5 | 9.8 | 5 | 9.4 |  |
| **Has the COVID-19 health situation changed your treatment plan?** |  |  |  |  | 0.80 |
| No | 51 | 70.8 | 41 | 67.2 |  |
| Yes | 11 | 15.3 | 12 | 19.7 |  |
| Unsure | 10 | 13.9 | 8 | 13.1 |  |
| **What role did you play in making that decision?** |  |  |  |  | 0.41 |
| I made the decision with little or no input from my doctor | 5 | 8.6 | 4 | 7.0 |  |
| I made the decision after seriously considering my doctor's opinion | 3 | 5.2 | 6 | 10.5 |  |
| My doctor and I share responsibility for the decision together | 32 | 55.2 | 31 | 54.4 |  |
| My doctor made the decision after seriously considering my opinion | 4 | 6.9 | 8 | 14.0 |  |
| My doctor made the decision about my treatment with little or no input from me | 14 | 24.1 | 8 | 14.0 |  |
| **How did your treatment plan change? (select all that apply)** |  |  |  |  |  |
| Delayed surgery | 3 | 4.2 | 2 | 3.3 | 1.00 |
| Delayed chemotherapy infusion | 2 | 2.8 | 7 | 11.5 | 0.08 |
| Delayed radiation | 1 | 1.4 | 0 | 0.0 | 1.00 |
| Stopped chemotherapy earlier than planned | 0 | 0.0 | 2 | 3.3 | 0.21 |
| **What do you believe contributed to the decision to change your treatment plan? (select all that apply)** |  |  |  |  |  |
| Concern about my COVID-10 exposure risk | 7 | 63.6 | 12 | 100.0 | 0.04 |
| Concern about availability of hospital beds and supplies | 2 | 18.2 | 5 | 41.7 | 0.37 |
| Concern about the availability of donated blood | 2 | 18.2 | 1 | 8.3 | 0.59 |
| Hospital/clinic rules related to COVID-19 | 5 | 45.5 | 4 | 33.3 | 0.68 |
| Professional medical organization recommendations | 0 | 0.0 | 1 | 8.3 | 1.00 |
| Strict visitor policy | 3 | 27.3 | 5 | 41.7 | 0.67 |
| Transportation concerns | 2 | 18.2 | 2 | 16.7 | 1.00 |

Almost all participants noted a change to at least some telehealth from in-person clinical visits. More than half of participants stated that decision-making was shared between the patient and physician; 7.2% reported that the decision was made “with little or no input from my doctor”, and conversely 19.2% reported that the decision was made “with little or no input from me.” The most commonly specified form of treatment modification was delay of chemotherapy infusion (38.5%), followed by delayed appointments or imaging. Concern about risk of exposure to COVID-19 was the most commonly cited reason for any changes (80.8%); other reasons included hospital or clinic policies (38.5%) or strict visitor policies (30.8%).

## DISCUSSION

Patients with cancer undergoing active treatment had the highest level of concern of getting infected by SARS-CoV-2. More than 90% in active treatment feared having a severe manifestation of the infection. This same population reported the highest level of family distress caused by the COVID-19 pandemic, indicating that the influence of related anxiety extended beyond just individual concern. Nearly 20% of participants reported that treatment changes during the pandemic were done “with little or no input” from them, but rather the logistical changes of cancer care were initiated exclusively or nearly exclusively by their treatment team.

The full impact of the COVID-19 pandemic on cancer patients may not be known for years, if not decades. In the immediate short-term, the current situation is rapidly altering the landscape of acceptable modifications to treatment in order to most effectively balance safety with efficacy of treatment. The partnership between patient and physician is integral and crucial to achieving this goal. The major concerns noted by respondents to our survey align with those reported on April 17, 2020 in the American Cancer Society’s (ACS) Findings Summary from its recent advocacy survey on the pandemic’s impact on cancer patients and survivors^11^. Key findings from that survey included 50% of cancer patients and survivors reporting impact on their healthcare during this time, with 27% of all patients who had been undergoing active treatment reporting some delay. Similar to our survey, 40% of ACS respondents undergoing active treatment expressed acute concerns about effects on their cancer-directed therapy plans^11^.

Our survey identifies some important gaps in knowledge. Specifically, what risks are posed to those that have a history of cancer but have completed treatment and are in less frequent contact with their care team? This group perceived their risk for contracting SARS-CoV-2 as being similar to, and notably lower than, those on active treatment. However, they also were concerned that they were at higher risk for developing severe disease. How susceptibility and disease pathogenesis are impacted by prior cancer therapy is a pressing research question, given the large number of survivors who have completed treatment and the accelerated aging and increased cardiovascular disease that often accompanies chemotherapeutic treatments.

One strength of this study was harnessing social media to quickly disseminate a research study in a relatively short period of time. However, this method led to disproportionate representation of cancer patients residing in Minnesota and the Midwest, thus limiting generalizability of study results to all cancer patients and the general population. The majority of respondents was skewed toward a specific (white and female) demographic; increased racial and sex diversity in respondents may have provided different results. Furthermore, the heterogeneity of cancer types represented, with inherent differences in treatment strategies and effect of treatment modifications and delays, impacts the ability to report specific alterations in treatment plans and the effect of specific management alterations on patient emotional state.

In summary, patients undergoing active treatment for cancer harbor concerns for short-term effects of the COVID-19 pandemic on the logistics and potential efficacy of ongoing cancer treatment, longer term effects, and overarching societal concerns that the population at large is not as concerned about the public health implications of the coronavirus. The healthcare landscape is quickly evolving to meet the needs of patients by attempting to balance safety with treatment efficacy. Over the long term, some of these adaptations, including virtual visits, may be permanently adopted by patients and healthcare systems as integral to overall cancer care. Regardless of the logistical approaches taken, partnership and adequate levels of communication between cancer care teams and patients with cancer are and will continue to remain critical during and for a long time following the current pandemic.

## Data Availability

Data are freely available.

